# Discordance Between Biomarker-Confirmed Antiretroviral Therapy and Self-Reported HIV Status Among People Living with HIV in Zambia and South Africa: A Secondary Analysis of HPTN 071 (PopART)

**DOI:** 10.64898/2026.07.22.26358720

**Authors:** Rita Nakalega, Dan Haines, Richard Hayes, Susan H. Eshleman, Helen Ayles, Peter Bock, Sian Floyd, Sarah Fidler, William Clarke, Yaw Agyei, Autumn Breaud, Brenda Mirembe Gati, Clemensia Nakabiito, Deborah Donnell

**Affiliations:** Makerere University-Johns Hopkins University (MU-JHU) Kampala, Uganda; College of Heath Sciences, Makerere University, Kampala, Uganda; Fred Hutchinson Cancer Center, Seattle WA, USA; Department of Infectious Disease Epidemiology, Faculty of Epidemiology and Population Health, London School of Hygiene &; Tropical Medicine, London, UK; Department of Pathology, Johns Hopkins University School of Medicine, Baltimore, Maryland, USA; Zambart University, Lusaka, Zambia; Desmond Tutu TB Centre, Department of Paediatrics and Child Health, Faculty of Medicine and Health Sciences, Stellenbosch University, Stellenbosch, South Africa; Imperial College National Institute for Health Research Biomedical Research Centre, London, UK

**Keywords:** HIV, self-report, antiretroviral therapy (ART), Sub-Saharan Africa

## Abstract

**Background:** Misclassification of HIV status in population-based surveys remains a critical barrier to accurate surveillance and program evaluation. Self-reported HIV status may diverge from objective measures, particularly among individuals receiving antiretroviral therapy (ART). We used biomarker-confirmed antiretroviral (ARV) drug detection to assess the prevalence and correlates of discordance between self-reported HIV status and biologic evidence of HIV treatment among people living with HIV (PLHIV) in Zambia and South Africa.

**Methods:** We conducted a secondary analysis of the HPTN 071 (PopART) cluster-randomized trial. At the 24-month survey visit, participants underwent HIV testing and laboratory assessment for ARV drugs in plasma. We defined discordant self-report (hereafter “non-disclosure”) as reporting HIV-negative or unknown status among individuals with ARV drugs detected. We estimated the prevalence of non-disclosure, compared prevalence by study arm, and used modified Poisson regression to identify associated factors. We also examined whether non-disclosure was associated with viral suppression (<400 copies/mL).

**Results:** Among 3,240 PLHIV with ARV drugs detected, 552 (17.0%) did not report an HIV-positive status—indicating that nearly one in six individuals on ART were misclassified by self-report. Non-disclosure did not differ between intervention and control arms (adjusted relative risk [aRR]: 1.03; 95% CI: 0.67–1.58). Non-disclosure was more common among younger individuals (age 18–24 years: aRR 2.30; 95% CI: 1.66–3.19), men (aRR: 1.39; 95% CI: 1.07–1.79), and those in formal employment (aRR: 1.42; 95% CI: 1.06–1.90). Individuals reporting condomless sex at last encounter were also more likely not to disclose (aRR: 1.59; 95% CI: 1.31–1.92). Viral suppression was high overall (93.7%) and did not differ by disclosure status (aRR: 1.06; 95% CI: 0.74–1.52).

**Conclusion:** A substantial proportion of PLHIV receiving ART did not report a known HIV-positive status, highlighting important discordance between biomarker evidence and self-reported data. Despite high levels of viral suppression, these individuals remain “hidden” from routine surveillance, with implications for estimating HIV diagnosis and treatment coverage. Strategies that incorporate objective measures alongside self-report, and that address social and structural barriers to disclosure, are essential to improve the accuracy of HIV surveillance and guide effective public health responses.

## INTRODUCTION

Eastern and Southern Africa has the largest number of people living with HIV (PLHIV), estimated at over 20.8 million (1). Expansion of antiretroviral therapy (ART) programs in these regions has significantly improved health outcomes of PLHIV, resulting in viral suppression, enhanced quality of life, and prolonged survival (2). Beyond individual health benefits, ART plays a crucial role in HIV prevention, as viral suppression reduces transmission risk (3–5). By the end of 2023, approximately 84% of PLHIV aged 15 years in Eastern and Southern were receiving ART, underscoring the scale of treatment coverage globally (1).

Accurate determination of HIV status is fundamental to effective HIV surveillance, clinical care and program evaluation (6), facilitating timely ART initiation, improving adherence, and fostering social support (7). In population-based surveys, HIV status is often determined by self-report, however, discordant self-report (hereafter “non-disclosure”) of an HIV-positive status remains a major challenge and self-reported HIV status may misclassify true treatment status (8–12). Such discordance between self-reported HIV status and biologic evidence of ART use represents an important measurement challenge in HIV surveillance and population-based estimates of HIV burden and treatment coverage (13).

Non-disclosure of HIV-positive status remains a key contributor to this misclassification and has implications profound consequences for both individuals and public health outcomes (14). At the individual level, non-disclosure delays linkage to care, and increases non-adherence to ART due to challenges taking medication openly and consistently (15, 16), which hinder the proper management of opportunistic infections, leading to worse health outcomes and increased morbidity (17).From a public health perspective, undisclosed HIV status can lead to misclassification of newly diagnosed cases and distortion of epidemiological data used for resource planning and policy implementation (18–20). Such data inaccuracies contribute to systemic inefficiencies that place additional strain on already overburdened healthcare systems in sub-Saharan Africa (18).

Stigma, discrimination, and concerns about confidentiality remain key barriers to disclosure, contributing to poor re-engagement in care (21, 22). In addition, survey context may influence self-report, with non-disclosure more likely in household or couple-based interview settings where privacy is limited (23). This study leverages biomarker-confirmed antiretroviral (ARV) drug detection to identify discordance between self-reported HIV status and biologic evidence of ART use, thereby addressing misclassification of HIV status in population-based surveys.

HPTN 071 (PopART) was a large three-arm cluster-randomized trial conducted in urban and peri-urban communities in Zambia and South Africa (24). Prior data from this cohort indicated that about 37% of PLHIV that had ARVs in their blood had disclosed that not been on ART (25). We conducted a secondary analysis of PopART data to identify predictors of non-disclosure of HIV status, assess differences in disclosure between intervention and control arms, and examine its association with viral suppression. This analysis aims to generate evidence on determinants of discordance in a high-burden setting to inform strategies for improving HIV surveillance accuracy, strengthening care engagement, and supporting progress toward global HIV target.

## METHODS

### The parent study design

The PopART study was a three-arm, cluster-randomized, trial conducted between 2013 and 2018 in 12 Zambian and nine South African urban and peri-urban communities (24). The study was designed to assess the impact of two community-level combination prevention packages on population-level HIV incidence across diverse community settings. These prevention packages included universal home-based HIV testing, linkage to HIV care, and expanded ART provision (24). The study’s main objective was to evaluate the effects of these interventions on HIV incidence, with a particular focus on understanding how different approaches to HIV testing and ART initiation influenced outcomes.

The community-level intervention strategies were implemented in 14 of the 21 study communities, covering a population of approximately 800,000 people. The study communities were matched into triplets based on estimated HIV prevalence, geographic location, and baseline ART coverage. In each triplet, communities were randomly assigned to 1 of 3 study arms. In Arm A, communities received a combination prevention intervention that included door-to-door HIV counseling and testing with universal ART (ART at any CD4 cell count). In Arm B, communities received the same combination prevention intervention with ART provided according to local guidelines, which shifted towards universal ART irrespective of CD4 count during the trial. In Arm C (control arm), communities received standard-of-care HIV testing services and ART according to local guidelines (24). The study assessed outcomes via annual follow-up (at 12, 24, and 36 months) of a large representative population cohort of adults aged 18-44 living in the selected communities. Surveys were conducted at participants’ homes, where individuals were offered rapid HIV testing during the study’s baseline and follow-up visits. Participants were asked about their HIV status during each visit, through response to the question, “What was your last HIV test result?”. The response options included HIV-positive, HIV-negative, don’t know, or not tested (24).

### Laboratory methods

Laboratory-based HIV testing was conducted in real-time for all participants in the Population Cohort (PC) participants at each study visit. A single fourth-generation HIV assay was used for the initial HIV testing and which was performed at central laboratories in South Africa and Zambia. Additional confirmatory testing to establish HIV status was conducted at the HPTN Laboratory Center (LC) in Baltimore, Maryland, USA. Viral load quantification (HIV viral load measurement) was performed using the Abbott RealTime HIV-1 assay (Abbott Molecular Inc., Des Plaines, IL), with a lower limit of detection of 400 HIV RNA copies/mL (24). Plasma samples were tested for ARV drugs using a qualitative liquid chromatography–high-resolution mass spectrometry assay. The method used identified 22 ARV drugs across five drug classes with detection limits ranging from 5 to 150 ng/mL, depending on the drug (25).

### Data Collection

For this secondary data analysis, we used ARV drug detection as a proxy for knowledge of a positive HIV status at the 24-month visit (i.e., we presumed that PLHIV who were taking ARV drugs were aware of their HIV-positive status). Our primary outcome variable was non-disclosure, defined as a participant reporting that he/she was HIV-uninfected or didn’t know his/her HIV status. We interpret this measure as encompassing both intentional non-disclosure and discordance between biologic evidence and self-reported status. The analysis included all participants who completed the 24-month (PC24) visit, were determined to be positive for HIV, and had ARV drugs detected in their plasma. The de-identified HPTN 071 (PopART) study dataset was accessed for this analysis on 15 June 2024. The dataset contained no personal identifiers, and the authors did not have access to information that could identify individual participants during or after data collection.

### Data analysis

Descriptive statistics summarized baseline characteristics including demographics, sexual risk behaviors, alcohol use, and self-reported HIV status (disclosed HIV positive, non-disclosed HIV positive). Demographic variables included age, gender, type of employment education status, household size, marital status and circumcision for men. Sexual risk factors included the number of sexual partners and occurrence of unprotected sex in the past 12 months. Alcohol use was assessed via the AUDIT score, reported both continuously and categorically.

Univariate modified Poisson regression models evaluated associations between baseline factors and HIV disclosure at PC24, presenting relative risks (RR) with 95% confidence intervals (CI). Demographic variables and sexual risk variables were included as predictors. Variables with a p-value <0.10 in univariate analysis were included in the multivariable regression model to estimate adjusted relative risks with 95% confidence intervals for non-disclosure.

To assess whether the intervention affected non-disclosure, an adjusted two-stage analysis was conducted to assess effects at the cluster level, following the study primary analysis approach (26). First, logistic regression modeled the outcome disclosure status against the matched (triplet), age-group, sex, and their interaction, excluding study arm. Predicted non-disclosure proportions for each community were calculated, adjusted for these factors. Observed-to-predicted risk ratios (ratio-residuals) for each community were computed via a Logistic model. The intervention effect was estimated by the mean difference in paired log ratio-residuals, with statistical significance assessed using two-way ANOVA (triplet and arm) and pairwise t-tests. Note that because the HIV-testing approach was the same for Arm A and B, these arms were both assessed as the same intervention for the disclosure outcome. P-values and confidence intervals were derived from these analyses.

Finally, we examine whether disclosure was a predictor for the outcome viral suppression, defined as HIV RNA <400 copies/mL at PC24. A multivariable modified Poisson regression model was used to examine the association between disclosure status and viral suppression, adjusting for potential confounders, including age, sex, marital status, form of employment, alcohol use and household size. Community was included as a fixed effect to account for contextual differences across communities.

## Results

A total of 21,678 participants completed the PC24 visit in the PopART trial. Of these, 4,902 tested HIV positive, and 3,240 had ARV drugs detected in their plasma were included in this analysis. Among PLHIV and taking ARV drugs, 552 (17.0%) did not disclose their HIV-positive status, reporting that they were HIV-negative or unaware of their HIV status.

Non-disclosure varied widely across communities from 5.3% to 43.1%. When summarized at the community level using the arithmetic mean of community-specific proportions, the average prevalence of non-disclosure was 17.9% in the intervention arms (Arms A and B combined) and 18.7% in the non-intervention arm (Arm C). The unadjusted and adjusted analyses comparing the disclosure proportion in Arms A and B compared to Arm C showed no statistically significant difference in non-disclosure rates associated with the PopART intervention (aRR; 1.03; 95% CI: 0.67–1.58; p = 0.90) **(Table 1).**

**Table 1:** Non-Disclosure of HIV Status by Study Arm and Triplet with Relative Risks and Adjusted Relative Risks by Subgroups at PC24.

| Group/Subgroup | Arm A Non-Disclosure, n/N (%) | Arm B Non-Disclosure, n/N (%) | Arm C Non-Disclosure, n/N (%) |
| --- | --- | --- | --- |
| **Overall** | 217 / 1070 (20.3%) | 163 / 1131 (14.4%) | 172 / 1039 (16.6%) |
| **Triplet 1** | 11 / 93 (11.8%) | 11 / 92 (12.0%) | 10 / 188 (5.3%) |
| **Triplet 2** | 17 / 149 (11.4%) | 38 / 290 (13.1%) | 9 / 122 (7.4%) |
| **Triplet 3** | 8 / 130 (6.2%) | 10 / 125 (8.0%) | 14 / 139 (10.1%) |
| **Triplet 4** | 38 / 223 (17.0%) | 28 / 171 (16.4%) | 25 / 157 (15.9%) |
| **Triplet 5** | 71 / 222 (32.0%) | 38 / 273 (13.9%) | 26 / 180 (14.4%) |
| **Triplet 6** | 52 / 185 (28.1%) | 10 / 115 (8.7%) | 85 / 246 (34.6%) |
| **Triplet 7** | 20 / 68 (29.4%) | 28 / 65 (43.1%) | 3 / 7 (42.9%) |
| Comparison | RR (95% CI), p-value | Adjusted RR (95% CI), p-value |  |
| Overall | 1.02 (0.66–1.58), 0.93 | 1.03 (0.67–1.58), 0.90 |  |
| Males | 0.91 (0.59–1.40), 0.63 | 0.85 (0.51–1.42), 0.50 |  |
| Females | 1.05 (0.65–1.69), 0.85 | 1.05 (0.65–1.69), 0.85 |  |
| Young (18–29) | 0.96 (0.44–2.12), 0.92 | 0.93 (0.38–2.26), 0.87 |  |
| Older (30+) | 1.05 (0.70–1.58), 0.80 | 1.05 (0.70–1.58), 0.80 |  |
| Young Males | 0.93 (0.64–1.36), 0.70 |  |  |
| Young Females | 1.00 (0.42–2.38), 0.99 | 0.97 (0.37–2.54), 0.94 |  |
| Older Males | 0.93 (0.58–1.50), 0.74 | 0.90 (0.47–1.69), 0.71 |  |
| Older Females | 1.09 (0.69–1.75), 0.68 | 1.09 (0.69–1.75), 0.68 |  |

Non-disclosers were younger (mean age 33.7 vs. 35.1 for those who disclosed); 12.3% of non-disclosers were aged 18–24 years compared to 6.1% in the disclosers. A higher proportion of non-disclosers were male (15.4% vs. 11.3%) and never married (47.3% vs. 30.3%). Formal full-time employment was more common among non-disclosers (10.0% vs. 7.8%). Fewer non-disclosers reported condom use at last sex (35.7% vs. 44.3%) (**Table 2**). Younger age was strongly associated with non-disclosure, with non-disclosure decreasing with increasing age. Participants aged 18–24 years had the highest risk of non-disclosure compared with those aged 40 years and older (aRR = 2.30; 95% CI: 1.66–3.19; p < 0.001), and those aged 25–29 years also had an increased risk (aRR = 1.37; 95% CI: 1.01–1.86; p = 0.042). Males were more likely than females not to disclose their HIV status (aRR = 1.39; 95% CI: 1.07–1.79; p = 0.013). Employment category influenced non-disclosure; participants in full-time formal employment had a higher risk (aRR = 1.42; 95% CI: 1.06–1.90; p = 0.020), while self-employed/other participants had a non-significant trend toward higher risk (aRR = 1.36; 95% CI: 0.97–1.92; p = 0.073). Not using a condom at the last sexual encounter in the past 12 months was associated with a higher risk of non-disclosure (aRR = 1.59; 95% CI: 1.31– 1.92; p < 0.001). Marital status, household size, number of sex partners, alcohol use, and education level were not significant in univariate analyses and were therefore excluded from the adjusted model. **(Table 3).**

**Table 2:** Baseline Characteristics of Participants by disclosure status.

| Characteristic | Overall (N = 3,240) | Disclosed HIV+ (N = 2,688) | Did not disclose HIV+ (N = 552) |
| --- | --- | --- | --- |
| **Age (years)** |  |  |  |
| 18–24 | 233 (7.2%) | 165 (6.1%) | 68 (12.3%) |
| 25–29 | 470 (14.5%) | 378 (14.1%) | 92 (16.7%) |
| 30–34 | 797 (24.6%) | 666 (24.8%) | 131 (23.7%) |
| 35–39 | 839 (25.9%) | 713 (26.5%) | 126 (22.8%) |
| 40+ | 901 (27.8%) | 766 (28.5%) | 135 (24.5%) |
| Mean (SD) | 34.9 (6.44) | 35.1 (6.31) | 33.7 (6.96) |
| Min, Max | 20, 46 | 20, 46 | 20, 46 |
| **Reported Gender** |  |  |  |
| Male | 390 (12%) | 305 (11.3%) | 85 (15.4%) |
| Female | 2,850 (88%) | 2,383 (88.7%) | 467 (84.6%) |
| **Type of Employment** |  |  |  |
| Unemployed / not working | 2,393 (73.9%) | 1,998 (74.3%) | 395 (71.6%) |
| Informal / irregular | 208 (6.4%) | 166 (6.2%) | 42 (7.6%) |
| Formal | 266 (8.2%) | 211 (7.8%) | 55 (10.0%) |
| Self-employed / other | 373 (11.5%) | 313 (11.6%) | 60 (10.9%) |
| **Marital Status** |  |  |  |
| Married / living as married | 1,537 (47.4%) | 1,330 (49.5%) | 207 (37.5%) |
| Never married | 1,075 (33.2%) | 814 (30.3%) | 261 (47.3%) |
| Divorced / separated | 380 (11.7%) | 326 (12.1%) | 54 (9.8%) |
| Widowed | 224 (6.9%) | 202 (7.5%) | 22 (4.0%) |
| Missing | 24 (0.7%) | 16 (0.6%) | 8 (1.4%) |
| **Number of sex partners (past 12 months)** |  |  |  |
| 0–1 | 2,945 (90.9%) | 2,453 (91.3%) | 492 (89.1%) |
| 2+ | 156 (4.8%) | 135 (5.0%) | 21 (3.8%) |
| Missing | 139 (4.3%) | 100 (3.7%) | 39 (7.1%) |
| Mean (SD) | 0.8 (0.91) | 0.9 (0.86) | 0.8 (1.11) |
| Min, Max | 0, 20 | 0, 20 | 0, 20 |
| **Condom use at last sex (past 12 months)** |  |  |  |
| No | 921 (28.4%) | 757 (28.2%) | 164 (29.7%) |
| Yes | 1,387 (42.8%) | 1,190 (44.3%) | 197 (35.7%) |
| Missing | 932 (28.8%) | 741 (27.6%) | 191 (34.6%) |
| **Alcohol use** |  |  |  |
| Never | 2,328 (71.9%) | 1,914 (71.2%) | 414 (75.0%) |
| Monthly or less | 500 (15.4%) | 424 (15.8%) | 76 (13.8%) |
| 2–4 times/month | 164 (5.1%) | 145 (5.4%) | 19 (3.4%) |
| 2–3 times/week | 113 (3.5%) | 96 (3.6%) | 17 (3.1%) |
| 4+ times/week | 64 (2.0%) | 57 (2.1%) | 7 (1.3%) |
| **Missing** | **71 (2.2%)** | **52 (1.9%)** | **19 (3.4%)** |
| **AUDIT score** |  |  |  |
| 0–7 | 2,735 (84.4%) | 2,253 (83.8%) | 482 (87.3%) |
| 8–15 | 217 (6.7%) | 189 (7.0%) | 28 (5.1%) |
| 16–19 | 39 (1.2%) | 34 (1.3%) | 5 (0.9%) |
| 20+ | 41 (1.3%) | 36 (1.3%) | 5 (0.9%) |
| Missing | 208 (6.4%) | 176 (6.5%) | 32 (5.8%) |
| Mean (SD) | 1.9 (4.40) | 1.9 (4.51) | 1.5 (3.85) |
| Min, Max | 0, 36 | 0, 36 | 0, 25 |
| **Household size** |  |  |  |
| 1 | 1,172 (36.2%) | 976 (36.3%) | 196 (35.5%) |
| 2–4 | 1,524 (47.0%) | 1,243 (46.2%) | 281 (50.9%) |
| 5+ | 528 (16.3%) | 459 (17.1%) | 69 (12.5%) |
| Missing | 16 (0.5%) | 10 (0.4%) | 6 (1.1%) |
| **Education Status** |  |  |  |
| None | 61 (1.9%) | 51 (1.9%) | 10 (1.8%) |
| Some / all primary | 1,038 (32.0%) | 902 (33.6%) | 136 (24.6%) |
| Some / all high school | 1,984 (61.2%) | 1,609 (59.9%) | 375 (67.9%) |
| College / university | 117 (3.6%) | 99 (3.7%) | 18 (3.3%) |
| Other | 11 (0.3%) | 7 (0.3%) | 4 (0.7%) |
| **Missing** | **29 (0.9%)** | **20 (0.7%)** | **9 (1.6%)** |
| Circumcision (men only) |  |  |  |
| No | 208 (53.3%) | 177 (58.0%) | 31 (36.5%) |
| Yes | 177 (45.4%) | 125 (41%) | 52 (61.2%) |
| Missing | 5 (1.3%) | 3 (1%) | 2 (2.4%) |

**Table 3.** Predictors of non-disclosure.

| **Characteristic** | **Univariate RR (95% CI)** | **p-value** | **Multivariate RR (95% CI)** | **p-value** |
| --- | --- | --- | --- | --- |
| **Age (years)** |  | <0.001 |  | <0.001 |
| 18–24 | 2.11 (1.66, 2.67) |  | 2.30 (1.66, 3.19) |  |
| 25–29 | 1.30 (1.03, 1.64) |  | 1.37 (1.01, 1.86) |  |
| 30–34 | 1.08 (0.87, 1.33) |  | 1.18 (0.90, 1.55) |  |
| 35–39 | 1.00 (0.80, 1.24) |  | 0.94 (0.71, 1.24) |  |
| 40+ |  |  |  |  |
| **Gender** |  | 0.019 |  | 0.013 |
| Male | 1.30 (1.06, 1.58) |  | 1.39 (1.07, 1.79) |  |
| Female |  |  |  |  |
| **Employment** |  | 0.226 |  |  |
| Informal / irregular | 1.04 (0.79, 1.37) |  |  |  |
| Formal | 1.25 (0.98, 1.60) |  |  |  |
| Self-employed / other | 1.24 (0.94, 1.65) |  |  |  |
| Unemployed / not working |  |  |  |  |
| **Marital status** |  | 0.069 |  |  |
| Never married | 1.26 (1.04, 1.53) |  |  |  |
| Divorced / separated | 1.14 (0.86, 1.50) |  |  |  |
| Widowed | 0.87 (0.57, 1.31) |  |  |  |
| Married / living as married |  |  |  |  |
| **Number of sex partners (past 12 months)** |  | 0.639 |  |  |
| 0–1 | 1.10 (0.73, 1.65) |  |  |  |
| 2+ |  |  |  |  |
| **Condom use at last sex (past 12 months)** |  | <0.001 |  | <0.001 |
| Yes |  |  |  |  |
| No | 1.68 (1.39, 2.03) |  | 1.59 (1.31, 1.92) |  |
| **Alcohol use** |  | 0.569 |  |  |
| Never | 1.27 (0.64, 2.54) |  |  |  |
| Monthly or less | 1.35 (0.66, 2.78) |  |  |  |
| 2–4 times/month | 0.98 (0.44, 2.16) |  |  |  |
| 2–3 times/week | 1.37 (0.61, 3.07) |  |  |  |
| 4+ times/week |  |  |  |  |
| **AUDIT score** |  | 0.926 |  |  |
| 0–7 | 1.20 (0.54, 2.67) |  |  |  |
| 8–15 | 1.11 (0.47, 2.62) |  |  |  |
| 16–19 | 1.11 (0.35, 3.46) |  |  |  |
| 20+ |  |  |  |  |
| **Household size** |  | 0.24 |  |  |
| 2–4 | 0.92 (0.76, 1.10) |  |  |  |
| 5+ | 0.80 (0.61, 1.04) |  |  |  |
| 1 |  |  |  |  |
| **Education status** |  | 0.484 |  |  |
| Some / all primary | 0.72 (0.40, 1.30) |  |  |  |
| Some / all high school | 0.83 (0.46, 1.49) |  |  |  |
| College / university | 0.80 (0.39, 1.63) |  |  |  |
| Other | 1.34 (0.45, 3.93) |  |  |  |
| None |  |  |  |  |

At PC24, viral suppression among PLHIV on ART was high, with 3,036 of 3,240 (93.7%) participants virally suppressed. Viral suppression was similar among those who disclosed their HIV-positive status (2,522/2,688; 93.8%) and those who did not disclose (514/552; 93.1%) In adjusted analysis, there was no evidence of an association between HIV disclosure status and viral suppression; participants who did not disclose their HIV-positive status had a similar likelihood of viral suppression compared with those who disclosed (aRR = 1.06; 95% CI: 0.74– 1.52; p = 0.735) **(Table 4).**

**Table 4:** Correlates of viral suppression status at PC 24.

|  | **Virally Suppressed** | **Not Virally Suppressed** | **Relative Risk**<br>**(95% CI)** | **p-value** | **Relative Risk**<br>**(95% CI)** | **p-value** |
| --- | --- | --- | --- | --- | --- | --- |
| HIV disclosure |  |  |  | 0.735 |  | 0.934 |
| Disclosed HIV- positive | 2522 / 2688 (93.8%) | 166 / 2688 (6.2%) |  |  |  |  |
| Did not disclose HIV-positive | 514 / 552 (93.1%) | 38 / 552 (6.9%) | 1.06 (0.74, 1.52) |  | 0.98 (0.68, 1.42) |  |

## DISCUSSION

In this secondary analysis of data from PopART trial we examined the predictors of non-disclosure of HIV status among individuals who had ARV drugs detected in plasma at the 24- month visit. Our findings highlight persistent gaps in disclosure of HIV status among PLHIV in a home-based survey, particularly among younger participants, males, those in formal employment and those never married.

Our findings align with prior research from sub-Saharan Africa indicating widespread non-disclosure during HIV testing campaigns and household surveys (18, 27). In Mozambique, 16.1% of individuals who tested positive during a provider-initiated testing campaign had previously been diagnosed with HIV, but did not disclose their status to providers(18). A national survey done in 2021 in the same country found that 14.1% of participants with ARV drugs detected failed to self-report their HIV status (27). Mistreatment by healthcare providers was the most cited reason for non-disclosure, followed by the desire to confirm a perceived cure and efforts to re-engage in care (18).

Strikingly, the 2012 Kenya AIDS Indicator Survey reported an even higher non-disclosure rate of 51.5% (288/559) among participants with ARV drugs detected (28). Non-disclosure in surveys leads to underestimation of HIV prevalence and treatment coverage, ultimately distorting national HIV estimates and misguiding policy decisions (12, 14, 29). In the surveys done, HIV diagnosis was under reported by 112,000 persons and 180,0000 in Kenya and Mozambique respectively (27, 28) and ART coverage was under-reported by 131,000 persons (27). If 17% of people with ARVs are misclassified, like in this study population-level estimates of awareness of HIV status and ART coverage may be systematically biased. Under reporting of HIV-positive status leading to under estimation of HIV prevalence by over 20% was found to be similar in Africa and North America in a systematic review (12). This PopART trial including several others, indicate that combining ARV drug testing with self-reported HIV and ART history improves the accuracy of identifying individuals who are aware of their HIV-positive (8–11).

Non-disclosure was significantly associated with younger age. Individuals aged 18–29 were less likely to disclose compared to those 40 and above. This aligns with findings from Mozambique, where those aged 15–24 had higher non-disclosure rates (27). Younger people often face complex disclosure challenges linked to stigma, limited autonomy, and fear of negative reactions from peers and family (30, 31). Gender disparities were also notable; men were significantly less likely to disclose, reflecting broader patterns of lower healthcare engagement driven by masculine norms that discourage vulnerability (32, 33).

Employment status was another key factor. Those engaged in full-time formal employment were more likely to withhold their HIV status compared to unemployed individuals. This may reflect differences in socioeconomic status, as seen in Kenya, where wealthier individuals were more likely not to disclose their HIV status (28). However, findings from another study in South Africa did not show a significant association between wealth and disclosure(34). These variable results may suggest that the relationship between socioeconomic status and disclosure may be context-dependent and influenced by factors such as healthcare experiences or perceived stigma.

Additionally, condom use at last sex was positively associated with disclosure, which may reflect underlying risk perception or greater engagement with prevention behaviors, as shown in other studies were condom use was associated with disclosure (35). Alcohol use, particularly higher AUDIT scores, showed no association with non-disclosure in this study though literature suggests that factors such as stigma, impaired judgment, and addiction can hinder engagement in care and interfere with effective communication with healthcare providers(36). These findings highlight the need to consider behavioral patterns when designing interventions to support disclosure and treatment adherence.

Despite high viral suppression among PLHIV on ART, non-disclosure remains a clinical and public health concern. Our findings challenge the common assumption that non-disclosure reflects poor engagement in care, as most non-disclosing individuals were virologically suppressed (∼94%). This suggests that non-disclosure may not necessarily indicate treatment failure or disengagement but may represent a subgroup of successfully treated individuals who remain unrecognized within routine HIV care and surveillance systems. However, non-disclosure may still hinder the delivery of tailored support and integrated care(37), underscoring the need to address disclosure barriers even among individuals who are successfully treated.

The strength of this study lies in its use of data from a large, well-characterized cohort within the rigorously designed PopART cluster-randomized trial, which improves the generalizability of findings to urban and peri-urban settings in sub-Saharan Africa. The combination of objective ARV drug detection, self-reported HIV status, and rigorously confirmed HIV status in a centralized reference laboratory enhances the accuracy for measuring non-disclosure, minimizing misclassification that can occur with self-report alone. The PopART study intervention team was present in the community for 2-3 years, making regular visits to all households, which could have contributed to lower rates of non-disclosure relative to the Indicator in Kenya (17% vs 51.5%) (28). Additionally, the availability of detailed demographic and behavioral data supported robust multivariable analyses, while advanced statistical methods accounted for clustering and confounding factors.

Some limitations should be noted. ARV detection was conducted only at PC-24 and does not allow determination of duration of ART use or differentiation between newly initiated and long-term ART users, which may affect interpretation of discordance. Key factors such as stigma and mental health were not measured. Additionally, behavioral data were self-reported, collected two years earlier at baseline and may have changed by PC-24, introducing potential social desirability and recall bias. This temporal separation limits causal interpretation and may attenuate observed associations. Mixed-methods research could have been useful to disentangle intentional non-disclosure from survey misclassification and better characterize the factors driving discordance.

## CONCLUSION

In this large cohort of people with confirmed HIV who had ARV drugs detected in plasma, about one in six did not disclose a HIV-positive status. Non-disclosure was more common among younger individuals, men, participants in formal employment, and those who did not use condoms at last sex. These findings highlight the need for differentiated strategies that address stigma, gender norms, and social isolation. Interventions that tackle emotional, social, and structural barriers to disclosure are essential to improve care engagement, ensure accurate surveillance, and advance progress toward epidemic control in high-burden settings.

## Data Availability

The data analyzed in this study are available publicly from the Harvard Dataverse at https://dataverse.harvard.edu/dataverse/HPTN-071. HPTN Statistical and Data Management Center (SDMC),

## Acknowledgements

We would like to thank all PopART study participants, and the teams at the South Africa and Zambia study sites.

## Authors’ contributions

RH was the HPTN071 Protocol Chair; SF was the HPTN 071 Protocol Co-Chair; DD was the HPTN 071 Protocol Statistician; SHE was the HPTN 071 virologist, PB was the HPTN 071 South African Site Co-Principal and HA was the HPTN 071 Zambian Site PI. RH, SF, SHE, DD, PB, HA, SF, HA, YA, WB were involved in designing and conducting the parent study.

RN and DD contributed the conception and design of this secondary data analysis study and provided project oversight. Data preparation and analysis were performed by DH. Results interpretation was done by RN and DD. Statistical oversight was provided by DD. The first draft of the manuscript was written by RN, and all authors commented on and edited previous versions of the manuscript. All authors reviewed and approved the final version of the manuscript. The corresponding author had final responsibility for the decision to submit for publication.

## Ethical Considerations

The POPART study was approved by the Institutional Review Board (IRB) at each participating site in Zambia and South Africa. All study procedures were conducted in compliance with IRB approvals, as well as relevant country and local regulations. Written informed consent was obtained from all participants in either English or their preferred local language.

## Consent for publication

Not applicable.

